# Unleashing the Potential of ^4^He OPM-MEG: A Comparison with SQUID-MEG for Detecting Interictal Epileptic Activity

**DOI:** 10.1101/2025.05.07.25326414

**Authors:** Denis Schwartz, Jean-Michel Badier, Tjerk P. Gutteling, Sébastien Daligault, Etienne Labyt, Francesca Bonini, Julien Jung

## Abstract

**Objectives:** Conventional magnetoencephalography (MEG) based on superconducting quantum interference devices sensors (SQUIDs), are the only widely used MEG systems in both clinical and research settings. However, they have limitations that hinder their widespread deployment. Optically pumped magnetometers (OPMs) offer several advantages over SQUIDs, particularly for epilepsy studies: lightweight and flexible, OPMs can be integrated into adaptable motion-tolerant headsets, enabling recordings during seizures or natural head movements, and potentially enhancing the detection of interictal epileptiform discharges (IEDs). In the present study, we assess the capabilities of a 5-sensors MEG system with helium OPMs (^4^He-OPMs) in detecting IEDs.

**Methods:** First, we compare the performance of SQUID-MEG and ^4^He-OPM-MEG in a routine clinical setup with a group of 7 patients. Second, we perform combined intra-cerebral (SEEG) ^4^He-OPM-MEG and SQUID-MEG recordings in a single patient to demonstrate the ability of both systems to detect IEDs originating from deep brain structures.

**Results:** The key finding is that, even with a very limited number of sensors, the ^4^He-OPM-MEG prototype successfully captured interictal epileptic activity in most patients. This activity was clearly detectable and exhibited the characteristic morphology with strikingly similar time courses between ^4^He-OPM-MEG and SQUID-MEG signals. Using combined SEEG and OPM-MEG recordings, we obtained the first direct validation of the ability of ^4^He OPM sensors to record epileptic activities originating from deep structures.

**Significance:** These results strongly support the clinical adoption of a lightweight, high-sensitivity, whole-head ^4^He OPMs-MEG system, offering new perspectives for epilepsy diagnostics and beyond, and enabling the democratization and spread of MEG in clinical and research settings.

**Plain language summary:** Magnetoencephalography is a non-invasive neuroimaging technique that has been shown to improve surgical outcomes in epileptic patients. However, its use remains limited due to several constraints, which could be overcome by a new generation of sensors: the optically pumped magnetometers (OPMs). Here, we validate the ability of OPM sensors to record epileptic brain activity in a regular clinical setup and thanks to simultaneous intracerebral recordings. These sensors open new venues for the widespread application of magnetoencephalography in the management of epilepsy and other neurological diseases, as well as for fundamental neuroscience.

**Key points:**

- Epileptic abnormalities are detected by ^4^He OPMs as well as with classical magnetoencephalography.
- Hippocampal interictal activity can be detected by ^4^He OPMs as shown by simultaeous SEEG-OPM recordings.
- This results represents significant step towards the validation of OPM-MEG for epilepsy diagnosis and neuroscience research.

## Introduction

Magnetoencephalography (MEG) is a non-invasive brain imaging technique that measures the very weak magnetic fields generated by neuronal activity, providing millisecond-level temporal resolution for real-time observation of brain function^1^. In the presurgical work-up of focal epilepsy, MEG offers significant advantages, including the detection of interictal discharges and, occasionally, ictal activity^2^. Thanks to advanced source localization techniques, MEG achieves high precision in identifying interictal epileptic foci^3^, providing unique insights that enhance the assessment and management of focal drug-resistant epilepsy^4^. Indeed, by non-invasively mapping the epileptic networks, MEG can guide both surgery and the placement of depth electrodes when invasive recordings are required, ultimately contributing to better post-operative outcomes^4^.

Conventional MEG systems based on superconducting quantum interference devices (SQUID) sensors^1^, are the only widely used and validated magnetoencephalography systems in both clinical and research settings. However, they have limitations that hinder their widespread deployment. SQUID sensors operate at extremely low temperatures (around 4°K or -269°C) and require cryogenic cooling, which must be supplied by liquid helium, making these systems expensive and complex to maintain. This cooling also necessitates a rigid and bulky Dewar, which restricts its use. The thermal insulation also imposes a sensor-to-scalp distance of 2–5 cm, reducing signal strength and spatial resolution, especially in children. Another limitation is the immobility required of participants, as head movement relative to the sensors introduces artifacts and degrades sources localization. Combined, these factors limit the clinical applications of MEG-SQUID systems, making them less suitable for studying populations such as infants, young children, or patients with involuntary movements such as during seizures.

Optically pumped magnetometers (OPMs) offer several advantages over SQUIDs, particularly for epilepsy studies, due to their design and operational characteristics. OPMs are wearable MEG sensors that detect modulations in the magnetic field using a gas cell as the sensitive element and laser absorption to measure density fluctuations^5,6^. OPMs don’t need cryogenic cooling making them more cost-effective and easier to use. Placed directly on the scalp, just millimeters from the brain, they provide significantly higher signal amplitude—up to 8 times stronger—and improved spatial resolution compared to SQUIDs^7,8^. Lightweight and flexible, OPMs can be integrated into adaptable motion-tolerant headsets, enabling recordings during seizures or natural head movements, particularly in children^9^, and potentially enhancing the detection of interictal epileptiform discharges (IEDs). All these features thus position OPMs as a transformative advancement, likely to increase the accessibility, versatility, and clinical impact of MEG.

To date, most OPMs studies have utilized sensors based on alkali gases (usually Rubidium). These studies have demonstrated the excellent performances of these sensors for recording physiological brain activity^9^ and pathological brain activity especially in epileptic patients^10–14^. The present study employs a different generation of OPMs sensors, tri-axial helium-4 optically pumped magnetometers (^4^He-OPMs)^6^. These offer a wide dynamic range (>200 nT) and a large bandwidth (DC to 2 kHz) allowing the recording of the whole spectrum of brain activities and significant head movement tolerance without signal saturation in lightly shielded environments. While the ability of ^4^He-OPMs to record physiological brain activity has been demonstrated in healthy volunteers^15^, only two proof-of-concept studies have shown that these sensors can also record epileptic activity. In the first study^16^ from Feys et al., 4 Rb-OPMs and 4 ^4^He-OPMs were used to detect IEDs in a child with refractory focal epilepsy. Over 1,000 IEDs were detected by both types of OPMs, with independent component analysis (ICA) used to isolate them from noise. In the second study^17^, Badier et al. used simultaneous recordings with intracerebral EEG (SEEG), SQUID-MEG, and ^4^He-OPM-MEG in a single patient to assess epileptic activity, demonstrating that 4 ^4^He-OPM-MEG could detect IEDs with a higher signal-to-noise ratio (SNR) than SQUID-MEG.

In the present study, we aim to assess the capabilities of a 5-sensors MEG system with ^4^He-OPMs in detecting epileptic activity. First, we compare the performance of SQUID-MEG and ^4^He-OPM-MEG in a routine clinical setup with a group of 7 patients. Second, we perform combined SEEG ^4^He-OPM-MEG and SQUID-MEG recordings in a single patient to demonstrate the ability of both systems to detect IEDs originating from deep brain structures. This allow us to assess the extent to which both systems can capture activity from regions that are typically harder to reach with non-invasive techniques. Through this study, we aim to improve our understanding of the strengths and limitations of ^4^He-OPM-MEG in noninvasive exploration of epilepsy.

## Methods

### Participants

7 participants suffering from epilepsy (age range 22-48, mean age 29.2 ± 8.9 years) took part in the study at the Hospices Civils de Lyon (Lyon, France). They all had an intractable epilepsy of diverse aetiologies which are described in table 1. The main selection criterion was an interictal EEG showing abundant epileptic abnormalities, mainly spikes. All participants signed an informed consent form prior to participation. The study was approved by the regulatory and ethics national administration in France (Study approved under number 2020-A00634-35 by l’Agence National de Sécurité du Médicament et des Produis de Santé et le Comité de Protection des Personnes SUD EST IV) and the patients gave informed consent.

**TABLE 1:**
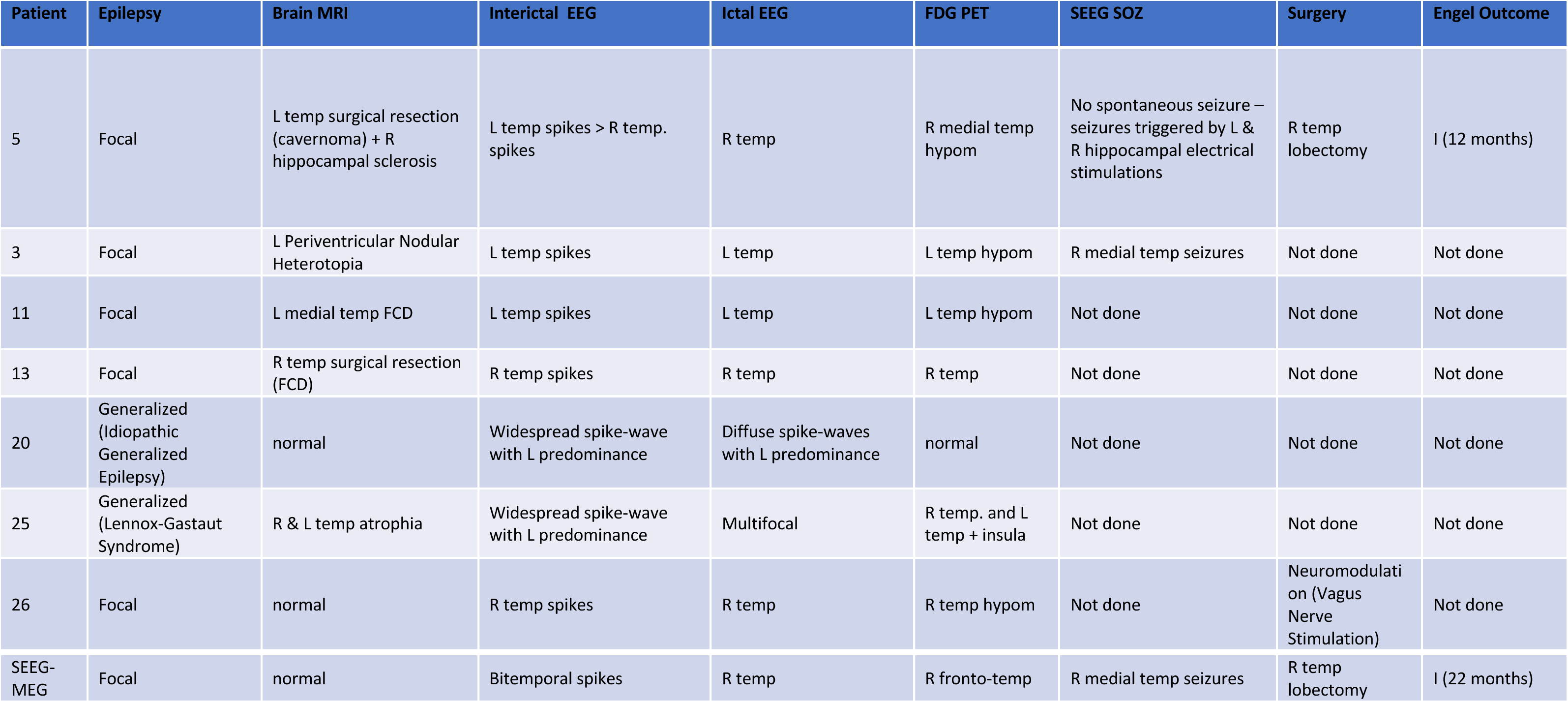
Patients: L: Left, R: Right, TEMP: Temporal, FCD: Focal Dysplasia, HYPOM: Hypometabolism, SOZ: Seizure Onset Zone.

The patient undergoing simultaneous SEEG-MEG recordings was hospitalized at La Timone Hospital (Marseille, France) for the invasive presurgical work-up of a drug-resistant non-lesional temporal lobe epilepsy. SEEG implantation, performed to assess laterality and the extent of the epileptogenic zone (EZ), included 14 depth electrodes, 11 on the right, exploring the temporal, insular and frontal regions, and 3 on the left, exploring the temporal region. The ethical approval was obtained at the regulatory and ethics national administration in France (Study approved under number 2020-A01830-39 by l’Agence National de Sécurité du Médicament et des Produis de Santé et le Comité de Protection des Personnes SUD EST IV) and the patient gave informed consent.

#### Data acquisition

##### SQUID-MEG and ^4^He OPM-MEG recordings

For the 7 patients undergoing non-invasive recordings, SQUID-MEG and ^4^He-OPM-MEG data were collected in a standard magnetically shielded room (MSR) (2 μ-metal, 1 copper layer, Vacuumschmelze, Hanau, Germany) without any active shielding.

SQUID-MEG data were acquired with a 275-channel CTF MEG system (CTF MEG Neuro Innovations, Inc., Coquitlam, Canada). The data were digitized at 2.4 kHz with a 1200Hz low-pass filter.

^4^He-OPM-MEG data were acquired using 5 triaxial ^4^He-OPMs sensors^6,18^ (MAG^4^Health, Grenoble, France) with one of the sensors serving as a reference sensor located on the top of the head, 10 cm above the scalp. The data were digitized at 11kHz.

Both recordings were done in up-right position on the chair used with the SQUID-MEG system. We systematically started with a SQUID-MEG resting state acquisition which lasted 30 minutes. An expert neurologist (JJ) visually analyzed the recorded signal to determine the scalp areas showing the higher occurrences of IEDs. The 4 ^4^He-OPMs sensors were then placed over these areas directly in contact with the scalp thanks to a semi-rigid helmet with 96 possible sensor positions. A wooden frame supported the ^4^He-OPMs sensors cables. The subjects were asked to minimize any head movement. The ^4^He-OPM-MEG resting state acquisition also lasted 30 minutes.

##### Simultaneous SQUID-MEG SEEG and ^4^He OPM-MEG SEEG recordings

For the patient undergoing SEEG, the simultaneous recordings were performed inside a similar 2-layer µ-metal magnetic shielded-room, in supine position. A first SQUID-MEG/SEEG recording session of 20 minutes at rest was followed by a comparable ^4^He-OPM-MEG/SEEG session. SQUID-MEG Signals were acquired with a 4D Neuroimaging™ 3600 whole head system at a sampling rate of 2034.51 Hz with a total of 248 axial magnetometers. The 4 triaxial

^4^He-OPMs sensors fixed on a rigid helmet were placed over the left central and temporal regions, in contact with the bandage covering the scalp. Placement of the sensors in this location was decided on the basis of IEDs on SEEG, which on that day were clearly more abundant in the left temporal regions than in the right. For more technical and methodological details on SEEG recording and simultaneous SEEG-MEG acquisition, see Badier et al, 2023^17^.

#### Data analysis

##### SQUID-MEG and 4He OPM-MEG data

All MEG data were analyzed using MNE-python^19^ (version 1.6.0). Analysis pipelines for SQUID-MEG and ^4^He-OPM-MEG were designed to optimize signal quality in terms of SNR for each modality.

SQUID-MEG denoising: We used the built-in CTF noise correction (3^rd^ gradient correction^20^) to remove any external interferences on the SQUID-MEG signals. The signals were then band-pass filtered between 3Hz and 70Hz. In patient 11, where the signal was heavily contaminated with dental artifact, the temporally extended signal space separation (tSSS) method^21^ was applied (window=120s, correlation=0.9). For further comparison with the ^4^He-OPM-MEG data, SQUID-MEG data was then restricted to the 4 sensors that were closest to the 4 ^4^He-OPMs sensors location in terms of Euclidian distance (See Figure S1).

^4^He-OPM-MEG denoising: The ^4^He-OPMs are triaxial sensors (one radial and two tangential axes) however the sensitivity on the second tangential axis is approximately a factor 4 lower than the other axes (∼200 ft/√Hz vs. <45 ft/√Hz). In this study we did not analyze the signal from this third axis to characterize the epileptic abnormalities however together with the signals from the reference sensor it was used to remove the low frequency artifacts contaminating the brain signals (see below).

We band-pass filtered the data between 6Hz and 70Hz, applied a notch filter at 50Hz and 60Hz and then the signal was epoched in 20s contiguous segments. To remove the low frequency artefact due mainly to subject head movement, the 3 axes of the reference channel data from the sensor and the second tangential axis of each of the brain channels were used (7 channels). These channels were low-pass filtered at 7Hz and used in a linear regression per trial and sensor to remove the low frequency noise from the 4 scalp sensors. After this step, epochs high amplitude events were marked and ignored in further processing (epochs removed: 7.0 ± 8.1). An ICA decomposition (fastica) was then applied. ICA components related to remaining gross non-neural artifacts were removed (components removed: 3.0 ± 1.9).

Upon visual inspection, an expert neurologist (JJ) marked all the epileptic abnormalities in both modalities. For both modalities and each abnormality, a SNR value was computed as follows: the maximum amplitude of the spike divided by the standard deviation of a baseline (20s of signal without abnormality).

The SNR values were compared through a t-test at the group level and the individual level. When necessary, we applied a Bonferroni correction to handle multiple comparisons.

##### Simultaneous SEEG/OPM-SQUID and SEEG/OPM-MEG

The 4 SQUID sensors closest to the 4 ^4^He-OPMs sensors were selected for analysis and for comparison with the magnetic activity recorded by the ^4^He-OPMs sensors. To synchronize recordings from different devices, triggers were sent to the two MEG systems (SQUID or ^4^He-OPMs) and SEEG. An in-house Matlab routine aligned the triggers and resampled SEEG data to match the MEG time frame, ensuring synchronization with minimal delay.

Noise compensation for SQUIDs was obtained by subtracting the contribution of the noise measured by the references (3 magnetometers and 9 gradiometer) for each sensor. ^4^He-OPMs lacked enough reference sensors, so noise reduction relied solely on filtering. As the head of the patient was kept immobile by the rigid helmet, this simple denoising was sufficient. All data, SQUID and ^4^He-OPMs, were band-pass filtered (2-70 Hz) with an additional 50 Hz notch filter.

The IEDs SNR were computed as the event’s maximum amplitude divided by the baseline standard deviation.

To compare SQUID and ^4^He-OPMs data, we performed an automatic extraction of IEDs after manually selecting events of interest from a montage consisting of a bipolar SEEG contact for each brain region explored. Similar patterns were found in both sessions using pattern recognition (Matlab’s findsignal function), and the events with the most similar activity on the SEEG selected contacts were retained and averaged together. A typical pattern on B’1-2 contacts (located in the hippocampus) was chosen for its reproducibility and frequency. For more details on SEEG and MEG data analysis, refer to Badier et al, 2023^17^.

## Results

### Comparisons between ^4^He-OPM-MEG and SQUID-MEG surface recordings

#### Group results

Overall, IEDs were detected in both modalities in 5 patients. 2 patients showed no IEDs in both modalities. 975 IEDs were detected with the SQUID-MEG (mean 195 ± 137) and 417 IEDs were detected with the ^4^He-OPM-MEG (mean 83 ± 97). As shown on Figure 1.A, at the group level, the SQUID-MEG SNRs (8.37 ± 6.54) were higher than the ^4^He-OPM-MEG SNR (6.72 ± 4.86) however the effect size was low (p < 0.01, Cohen’s distance (cd) = 0.27). In term of maximum spike amplitude (Figure 1.B), the ^4^He-OPM-MEG showed larger values, as expected (3.76 ± 2,81 pT), when compared to the SQUID-MEG (1.66 ± 0.66 pT) with a large effect size (p < 0.01, cd = 1.28, amplitude ratio ^4^He-OPM-MEG / SQUID-MEG = 2.3). ^4^He-OPM-MEG radial measurements showed slightly higher SNR (4.42 ± 2.68) when compared to tangential measurements (4.12 ± 2.36) with a very small effect size (p < 0.01, cd = 0.12, Figure 1.C). The maximum spike amplitude was higher for the radial direction (2.28 ± 1.85 pT) than the tangential direction (1.48 ± 1.17 pT) (p < 0.01, cd = 0.51, amplitude ratio radial / tangential = 1.5, Figure 1.D).

**Figure 1.**
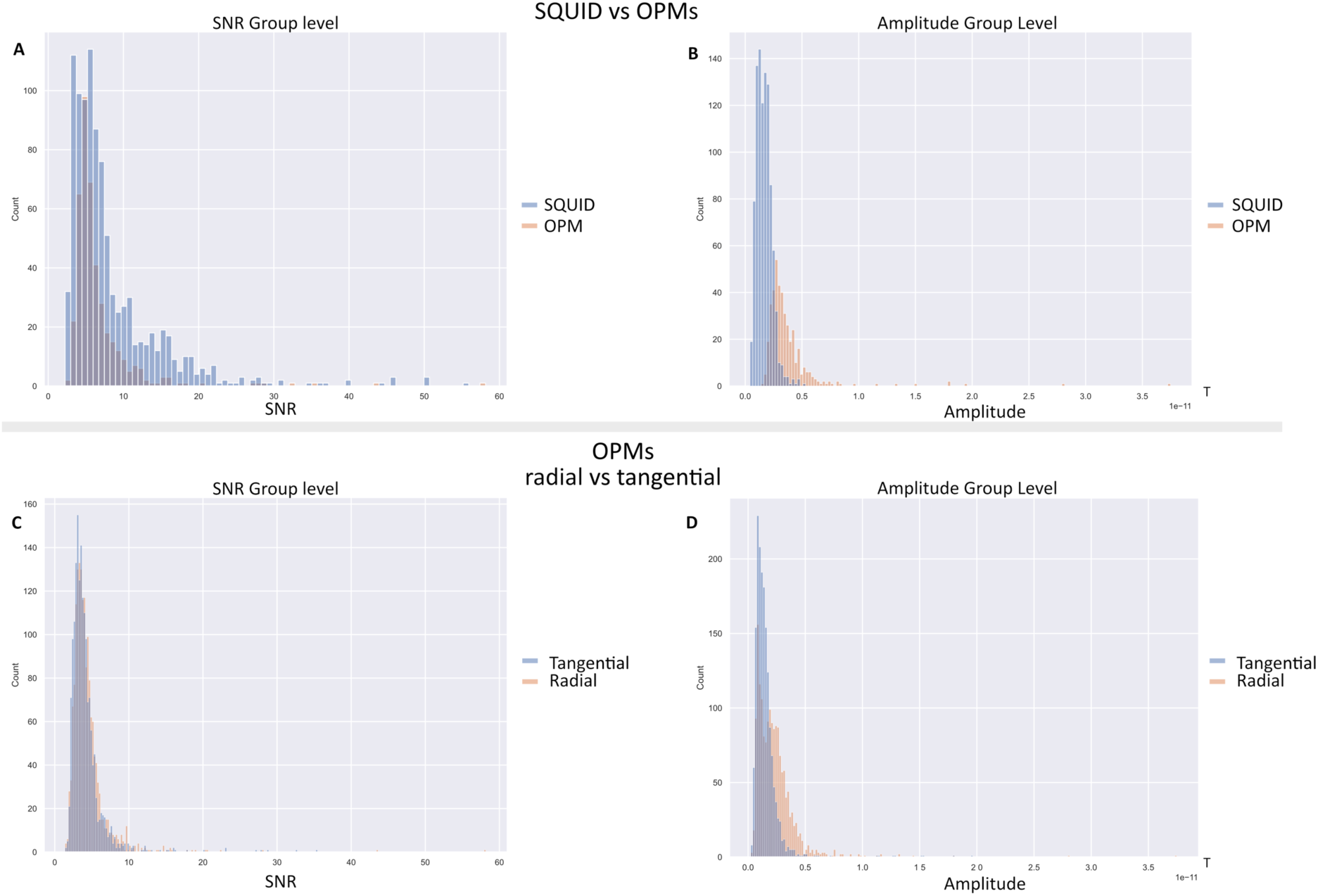
Group level results for SNR and amplitudes of detected IEDS - Top: SQUID-MEG versus ^4^He-OPM-MEG comparison A) Distribution of SNRs (SQUID-MEG in blue, and ^4^He-OPM-MEG in orange) – B) Distribution of epileptic spikes maximum amplitude (SQUID-MEG in blue, and ^4^He-OPM-MEG in orange). Bottom: ^4^He-OPM-MEG radial versus tangential comparison C) Distribution of SNRs (Tangential direction in blue, and radial direction in orange) – D) Distribution of epileptic spikes maximum amplitude (Tangential direction in blue, and radial direction in orange)

#### Individual results

The group results were largely confirmed at the individual level with SQUID-MEG SNRs being larger than ^4^He-OPM-MEG SNRs for 3 patients, although only one showed a large effect size (see Figure 2.A for patient 3, cd = 1.21) the two others being small (cd = 0.26 and 0.15) (See Figure 2.C for patient 20 – All individual results and statistical analysis are shown in tables S1 and S2 and Figure S2). Two patients showed ^4^He-OPM-MEG SNRs higher than SQUID-MEG SNRs, but the differences were not significant (p > 0.1). ^4^He-OPM-MEG signal amplitudes were significantly and systematically higher for all patients when compared to SQUID-MEG signal amplitudes (See Figure 2.B and 2.D for patient 3 and 20, p < 0.01, cd ranging from 1.5 to 6.5 and amplitude ratio ^4^He-OPM-MEG / SQUID-MEG ranging from 1.4 to 5.3). The increased level of noise with the ^4^He OPM-MEG is also very visible in most patients confirming the SNRs results at the group level. The ^4^He OPM-MEG radial component showed more prominent epileptic abnormalities. However, this was not systematic, as the tangential direction could also show clearer IEDs than the radial one, as illustrated in Figure 3.A (channel LT35) and in supplementary materials for patients 11 and 25. Finally, the morphologies of IEDs were most often similar in both SQUID-MEG and ^4^He-OPM-MEG recordings, as shown in Figure 3 for patients 3 and 20, and in supplementary materials for the other patients in Figure S3.

**Figure 2.**
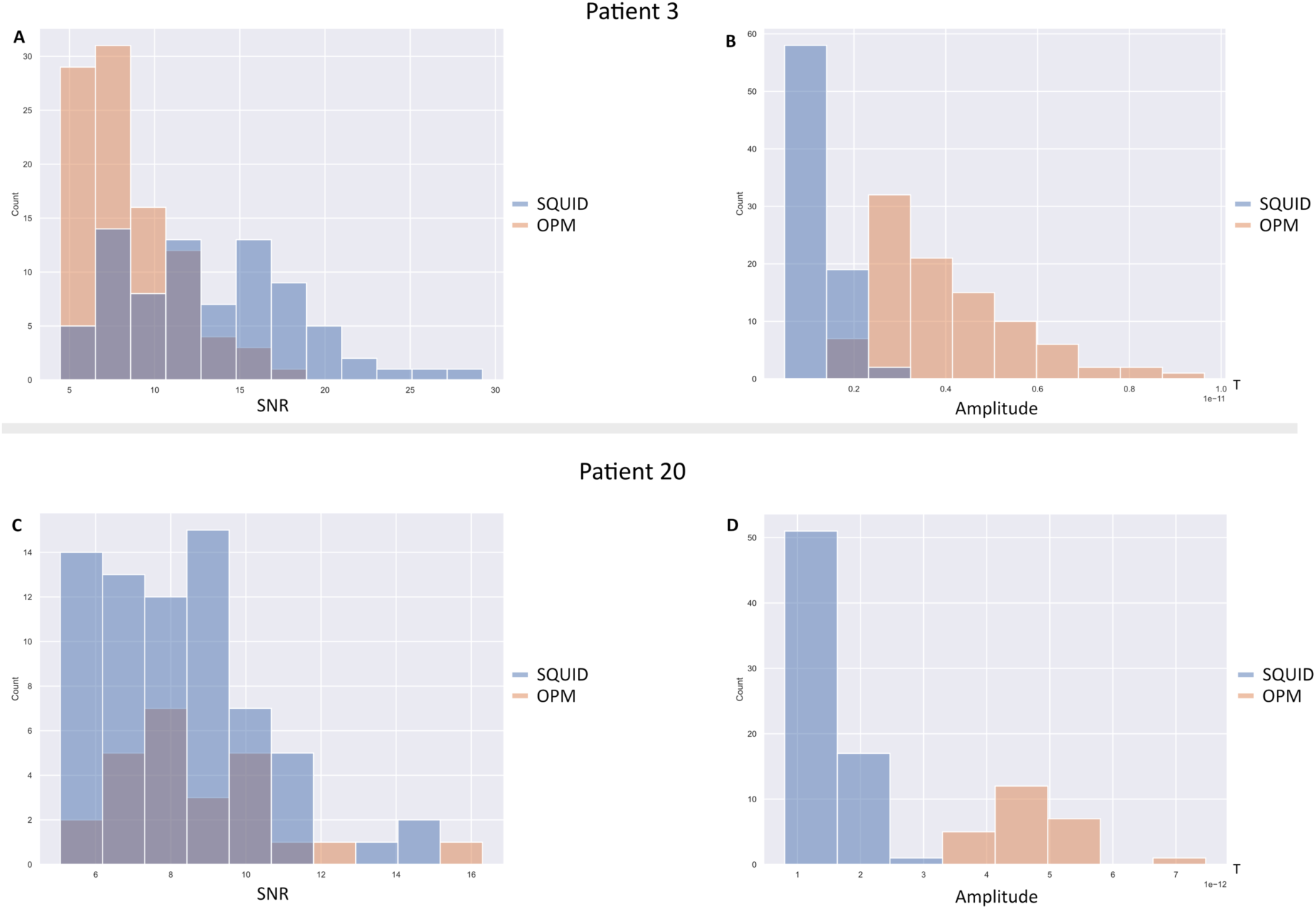
Individual results for SNR and amplitudes of detected IEDS for patients 3 (top) & 20 (bottom)– A, C) Distribution of SNRs (SQUID-MEG in blue, and ^4^He-OPM-MEG in orange) – B, D) Distribution of epileptic spikes maximum amplitude (SQUID-MEG in blue, and ^4^He-OPM-MEG in orange).

**Figure 3.**
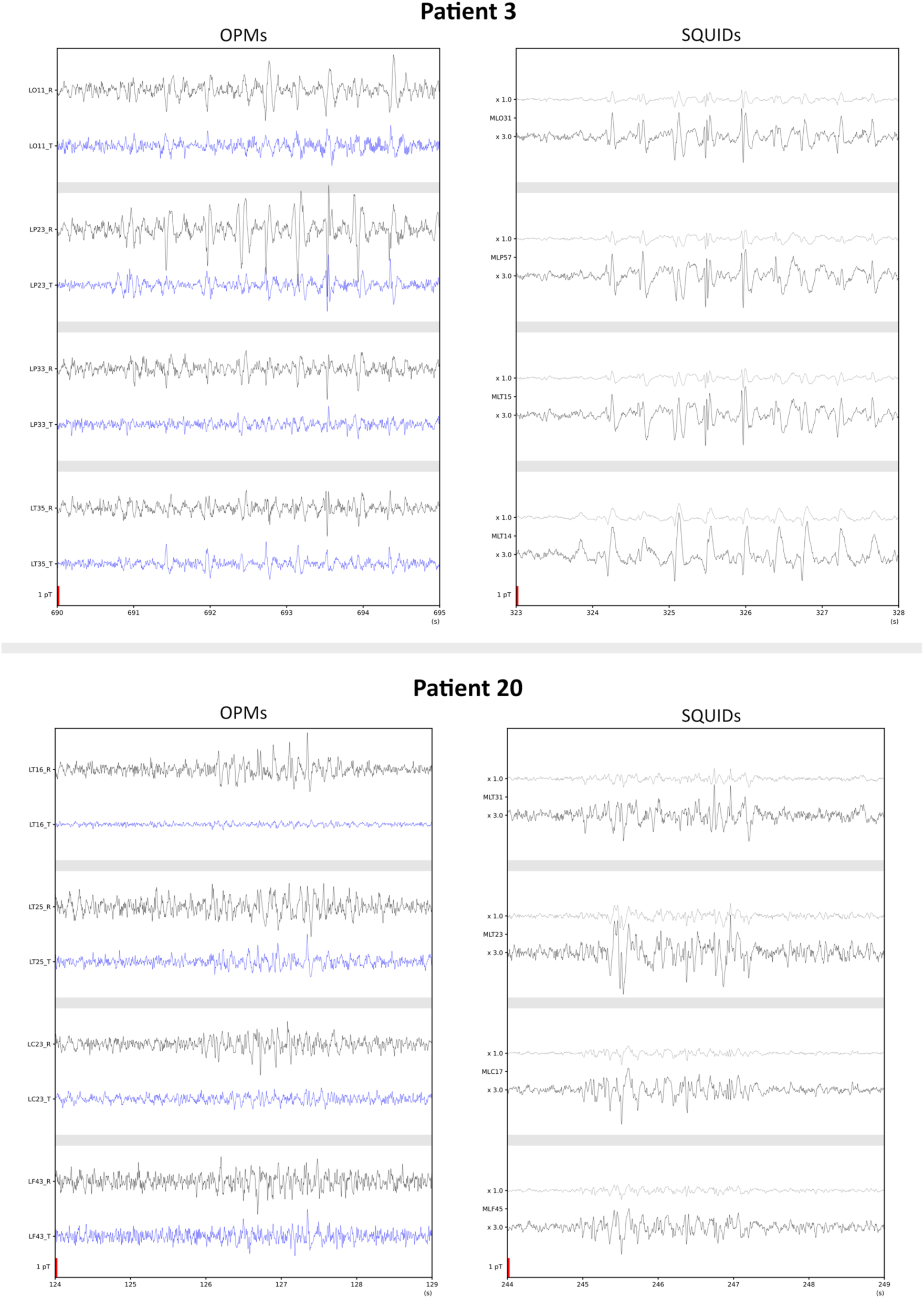
Patients 3 (top) & 20 (bottom) IEDs – For each patient the left panel shows the ^4^He-OPMs traces with for each sensor in black the radial axis and in blue the tangential axis. The right panel shows the closest SQUID channel for each ^4^He-OPMs sensors with, in light grey, the trace at the same scaling as the ^4^He-OPMs traces and in black the SQUID traces scaled by 3 for better comparison.

### SEEG results

While it was not possible to distinguish epileptic activity in the spontaneous MEG signal, it was possible to detect activity originating for deep brain structures after averaging signals recorded within the ^4^He-OPM-MEG session and within the SQUID-MEG session using SEEG spikes as triggers.

Averaged SQUID-MEG signals obtained from 23 epochs and averaged ^4^He-OPM-MEG signals from 20 epochs are showed in Figure 4, together with SEEG averaged IEDs.

**Figure 4.**
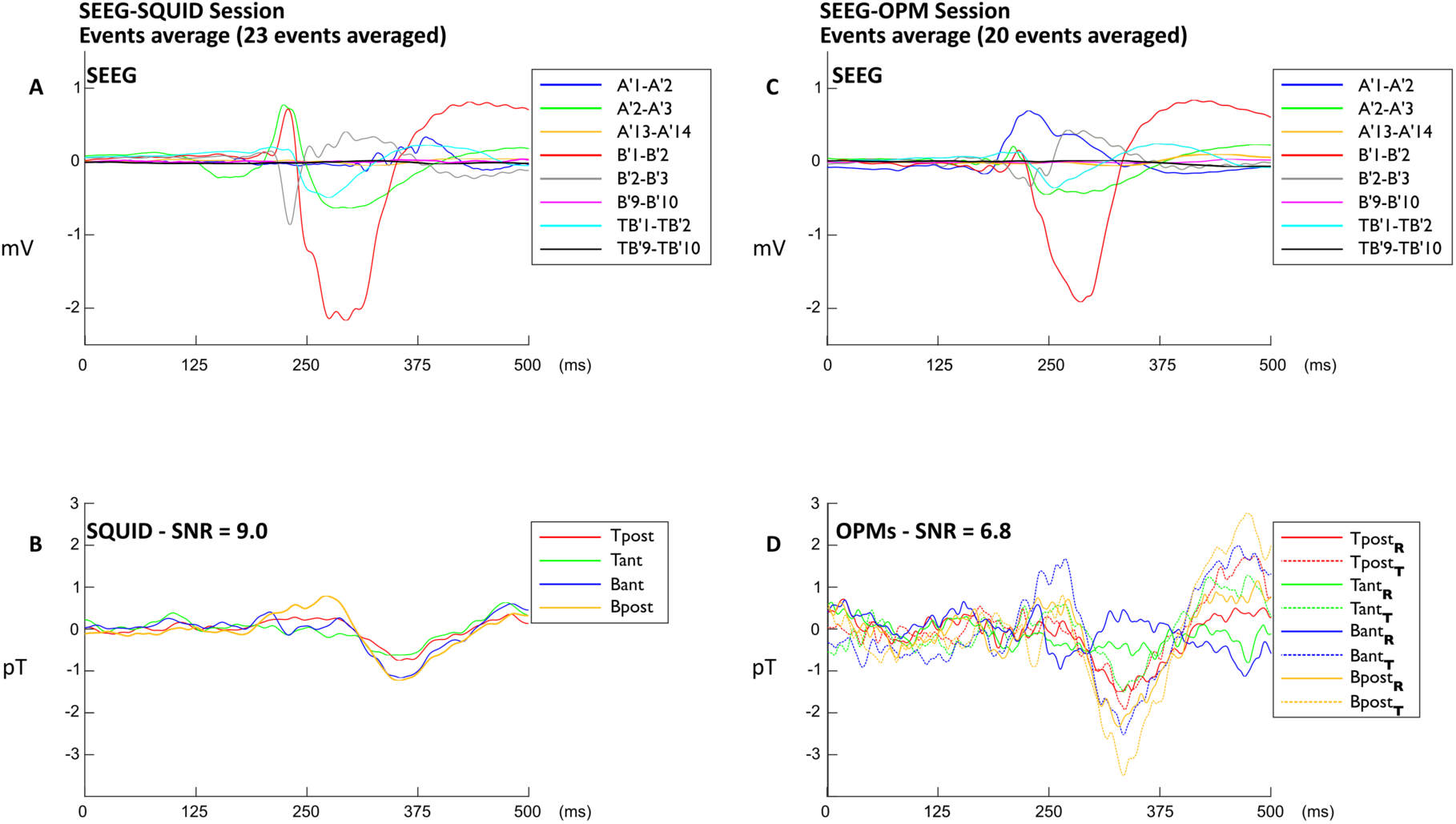
MEG-SEEG Patient: Averaged spikes. The left side of the figure presents the averaged signal (23 epochs) collected during the SQUID-MEG/SEEG simultaneous session. the right side presents the averaged signal (20 epochs) collected during the ^4^He-OPM-MEG/SEEG simultaneous session. The name of the SEEG electrodes and recording contacts are shown. Contacts range from 1 (deeper location) to 10 (most superficial location). Scales are identical between sessions for comparison. A, Bipolar averaged SEEG data for the SQUID session. The averaged spike involves the deepest recording contacts exploring the amygdala (A), the anterior part of the hippocampus (b) and the temporo-basal region (TB). No activity is noticeable in the areas explored by the most superficial contacts. C, Bipolar averaged SEEG for the ^4^He-OPM-MEG session, disclosing a very similar averaged spike arising from deep temporal structure. B, Simultaneous SQUID-MEG averaged data on the 4 SQUID sensors closest to the ^4^He-OPMs sensors. ‘T’ For top, ‘B for bottom’, ‘ant’ for anterior and ‘post’ for posterior. D, Simultaneous ^4^He-OPM-MEG averaged data on the four sensors. ‘T’ for Top sensors, ‘ant’ for anterior, ‘post’ for posterior, ‘T’ for tangential, ‘r’ for radial.

The SEEG average signals from both sessions are very similar, both in terms of time-courses and of signal amplitudes that remain consistent between the two sessions, except for a difference in the activity recorded in the amygdala (figure 4, top panels). No significant activity can be observed in SEEG contacts exploring superficial neocortical regions, indicating that recorded activity is primarily limited to medial temporal regions. Discrepancies in the timing between the peaks of activity across different SEEG contacts can be likely attributed to signal propagation within the deep temporal structures.

As expected, the amplitudes of the ^4^He-OPM-MEG signals are higher than SQUID-MEG signals. The SNR is slightly higher for SQUID-MEG (9.0) than for ^4^He-OPM-MEG (6.8). Interestingly, the distribution of magnetic fields differs between SQUID and ^4^He-OPM-MEG sensors. Indeed, while the time courses recorded on the 4 SQUID sensors are very similar, the time courses recorded on the 4 radial ^4^He-OPMs sensors shows a much greater variation in amplitude (Figure 4.D, solid lines). It is also worth noting the existence of responses of equivalent or greater amplitude on the tangential axes of ^4^He-OPMs sensors (Figure 4.D, dotted lines).

## Discussion

This study is the first to evaluate the performance of a novel prototype ^4^He-OPM-MEG device built with 5 ^4^He-OPMs sensors, compared to a conventional SQUID-MEG system for the detection of epileptic activity in a substantial patient cohort.

The key finding is that, even with a very limited number of sensors, the ^4^He-OPM-MEG prototype successfully captured IEDs in most patients. These IEDs were clearly detectable through standard visual analysis and exhibited the characteristic morphology of epileptic spikes with strikingly similar time courses between ^4^He-OPM-MEG and SQUID-MEG signals. Moreover, quantitative analysis comparing spikes recorded with the 4 ^4^He-OPMs sensors and their 4 nearest SQUID-MEG counterparts revealed SNRs in the same range, with a slight advantage (small effect size) for SQUID-MEG and higher signal amplitudes for the ^4^He-OPM-MEG. Lastly, using combined SEEG and ^4^He-OPMs recordings, we obtained the first direct validation of the ability of ^4^He-OPMs sensors to record epileptic activities originating from deep structures.

These findings highlight the strong potential of ^4^He-OPMs technology, paving the way for more accessible and flexible MEG systems in clinical and research applications.

To ensure the highest signal quality for each modality, we implemented preprocessing pipelines specifically optimized for SQUID-MEG and ^4^He-OPM-MEG. This tailored approach allowed us to fully leverage the strengths of each system. As shown in our previous study^15^, ^4^He-OPM-MEG signals, while more sensitive to motion artifacts due to sensors being directly attached to the patient’s head, benefited from advanced correction strategies. By using the additional axis of each sensor and reference sensors positioned 10 cm above the head, we effectively removed movement-related noise (between DC and 5 Hz). Meanwhile, SQUID-MEG signals naturally exhibited lower noise levels due to their gradiometer configuration and/or third-gradient correction^20^. In one case, the ability to apply tSSS on SQUID-MEG recordings proved advantageous in removing a dental artifact, which was less prominent in ^4^He-OPM-MEG data. By tailoring preprocessing to each system’s specific characteristics, we ensured optimal signal quality, maximizing the IEDs amplitude and SNR measurements. The recordings made during the simultaneous acquisition sessions probably benefited from the subject’s very limited mobility in the supine position and required only simple filtering to obtain interpretable traces.

While the spike detection rate was lower with ^4^He-OPM-MEG, it is remarkable that all patients who exhibited IEDs with SQUID-MEG also showed IEDs with ^4^He-OPM-MEG, confirming the reliability of this technology in detecting epileptic activity. Spike detection was performed across the entire MEG sensor array (275 channels) for SQUID-MEG, enabling a refined topographical analysis of candidate events, whereas only 4 sensors were used for ^4^He-OPM-MEG. The broader spatial coverage of the SQUID-MEG sensors likely contributed to the higher yield for detecting IEDs. Furthermore, because of the shorter source-sensor distance and lower sensitivity, the field maps of the ^4^He-OPMs sensors are much more focal, displaying higher spatial frequency compared to SQUID-MEG. Thus, if the ^4^He-OPMs are not optimally positioned, the IEDs may be less detectable or appear with reduced amplitude. Future studies evaluating whole-head ^4^He-OPM-MEG devices with full scalp coverage will improve spike detection performance during clinical recordings.

The sequencing of the experiment, with SQUID-MEG recordings always performed before ^4^He-OPM-MEG, may have also favored SQUID-MEG. Patients were generally able to sleep during the first session, but not during the second one after the ^4^He-OPM-MEG setup. This difference in sleep state could have significantly reduced the occurrence of abnormalities in the epileptic network^22^. Additionally, the reported detection rates of IEDs can vary in the literature, with studies showing both higher or lower detection rates with OPMs ^12,14^.

A direct and comprehensive comparison of diagnostic performance between SQUID-MEG and various OPM systems is not possible due to the significant difference in sensor count, with SQUID systems having over 200 sensors, whereas published OPMs studies use far fewer sensors. Nonetheless, we observed that ^4^He-OPM-MEG and SQUID-MEG detected IEDs with SNRs in the same range, though SQUID-MEG had a slight advantage, with a small effect size. These findings align with previous studies in adults^12,14,17^ using either ^4^He-OPMs or alkali OPMs. However, studies on epileptic children have reported significantly higher SNRs in OPMs compared to SQUID-MEG^11^, likely due to the closer proximity of OPMs to the brain, a benefit that is less pronounced in adults. As shown by several studies using ^4^He-OPM-MEG^15,17^ and alkali OPMs^12^, the signal amplitude was always larger for the ^4^He-OPM-MEG due to sensor proximity, the power ratio ranging from 2 and 5, consistent with prior research cited above. Notably, even higher ratios have been observed in children^11^.

As shown in previous studies using simulation^7,23,24^ and on healthy subjects^15^, the tangential measurement of the OPMs sensors is bringing complementary information to radial axis measurements. This was confirmed here on IEDs with equivalent SNRs at the group level between both axes. Furthermore, some IEDs were more clearly visible on the tangential axis than on the radial axis. Given their orientation, these channels could be more sensitive to neuronal populations not usually recorded with the radial components. It is conceivable that the detection and source localization accuracy of IEDs could be improved by incorporating information from multiple spatial directions.

The simultaneous MEG-SEEG recordings realized in a single patient, with SEEG serving as a reference and ground truth, have enabled a reliable comparison between SQUIDs and ^4^He-OPMs. The results further support the evidence that ^4^He-OPMs can detect IEDs at a level comparable to SQUIDs, with a lower sensitivity but a signal-to-noise ratio in the same order of magnitude. This confirms previous research demonstrating the ^4^He-OPMs correlate of intracerebral SEEG spikes originating from the temporal pole^17^. Moreover, although this concerns averaged rather than spontaneous activity, the results demonstrate that OPMs, like SQUIDs, can capture epileptic activity originating from deep structures such as the amygdala and hippocampus, opening new perspectives for the investigation of mesial temporal epilepsy, as suggested by a previous single case using alkali OPM^13^.

In the near future, the development of helmets with more sensors and sufficient head coverage is planned, enabling the application of signal processing techniques such as blind source separation to visualize spontaneous, non-averaged activity^25,26^. Additionally, the flexibility of OPMs sensors will allow their placement on the face and cheeks, reducing the distance to temporo-mesial structures and potentially enhancing their recording. This could make it possible to detect and localize spontaneous activity from deep sources—whether epileptic^25^ or physiological^26^—without the need for simultaneous SEEG, opening new possibilities for non-invasive cognitive mapping in normal and epileptic subjects.

Interestingly, ^4^He-OPMs signals obtained simultaneously to SEEG exhibit a considerable variability in time courses depending on the location of the sensors, indicating their ability to differentiate various components of brain activity. This advantage arises from their placement directly on the scalp, in close proximity to the brain, enabling signals with a higher spatial frequency to be recorded. Consequently, a greater number of sensors may be required to ensure comprehensive sampling of the magnetic field measured on the scalp, ultimately facilitating a more precise discrimination of the sources of recorded brain activity.

The limited number of ^4^He OPMs sensors recording brain activity is the major limitation of the study, precluding source localization of IEDs. However, the promising performance of the ^4^He-OPM-MEG system is particularly notable despite the constraints of the 5 sensors setup. Note that all acquisitions were done in a standard MSR without active shielding and that recent advancements have introduced a whole-head system with thin, flexible, and lightweight cabling and highly sensitive sensors (∼ 30 fT/√Hz noise floor)^24^.

A key advantage of the ^4^He-OPM-MEG system lies in the close proximity of its sensors to the scalp. This proximity enhances signal amplitude and partly compensates for the system’s lower intrinsic sensitivity for superficial sources. Additionally, the stability and wide dynamic range of these sensors enable long recording sessions, even with significant patient movement, facilitating the detection of IEDs.

Overall, this study builds upon previous findings in healthy subjects^15^ and epilepsy patients^17^, reinforcing the potential of ^4^He-OPM-MEG for clinical applications. These results strongly support the clinical adoption of a lightweight, high-sensitivity, whole-head OPM-MEG system, offering new perspectives for epilepsy diagnostics and beyond, and enabling the democratization and spread of magnetoencephalography in clinical and research settings.

## Acknowledgments

This study was supported by Agence Nationale de la Recherche Grants ANR-22-CE19-0012-01, Grant ANR-11-INBS-0006 to France Life Imaging network, and ANR-16-CONV-0002 to Institute of Language, Communication,and the Brain, and grant from Région Auvergne-Rhône-Alpes (Pack Ambition project: NEW_MEG). TG is supported by the Labex Cortex (ANR-11-LABX-0042).

We thank the nurses involved in the preparation and recording.

## Author Contributions

Conceptualization, D.S., E.L., J.J., J.M.B, F.B.; methodology, T.P.G., D.S., E.L., J.J., J.M.B, F.B.; software, T.P.G., D.S., J.M.B.; formal analysis—T.P.G., D.S., J.M.B; investigation, J.J., D.S., E.L., S.D. J.M.B., F.B.; data curation, D.S., J.M.B.; writing—original draft preparation, D.S., J.J., J.M.B., F.B.; writing—review and editing, T.P.G., S.D., E.L.; project administration—D.S. J.J., J.M.B.; funding acquisition, D.S., E.L. J.J., J.M.B., F.B.; All authors have read and agreed to the published version of the manuscript. All Authors confirm that they have read the Journal’s position on issues involved in ethical publication and affirm that this report is consistent with those guidelines.

## Institutional Review Board Statement

The study was approved by the regulatory and ethics national administration in France under number 2020-A01830-39 and 2020-A00634-35 by l’Agence National de Sécurité du Médicament et des Produis de Santé et le Comité de Protection des Personnes SUD EST IV.

Informed Consent Statement: Informed written consent was obtained from all subjects involved in the study.

## Data Availability Statement

Anonymized data supporting the results of this study are available from the corresponding author upon reasonable request and validation by regulatory and ethical bodies and subject consent.

