## Supplementary materials for "Unleashing the Potential of ^4^He OPM-MEG: A Comparison with SQUID-MEG for Detecting Interictal Epileptic Activity"

### Supporting Information: figures

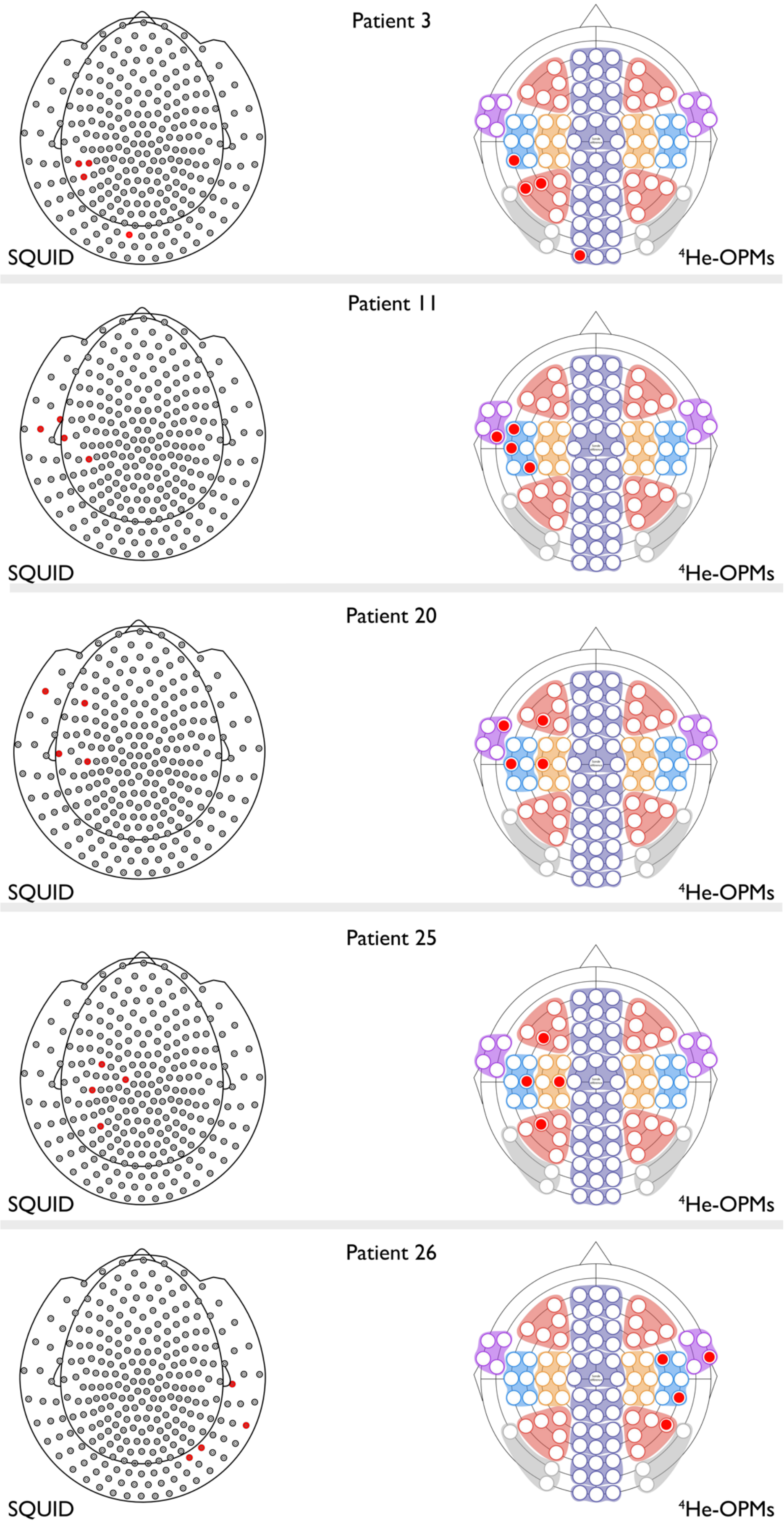

**Figure S1**: Sensors used in the analysis for SQUID-MEG (left) and OPM-MEG (right) for each patient

### Patient 3

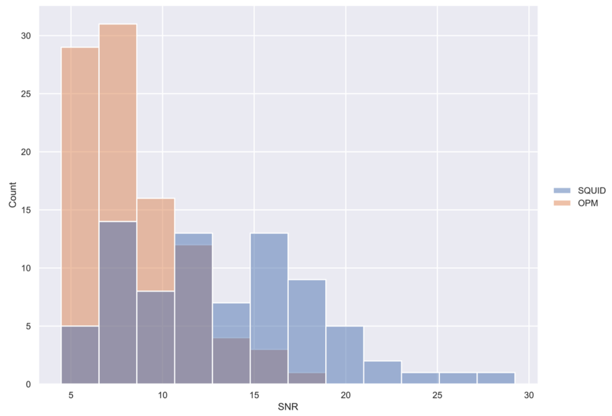

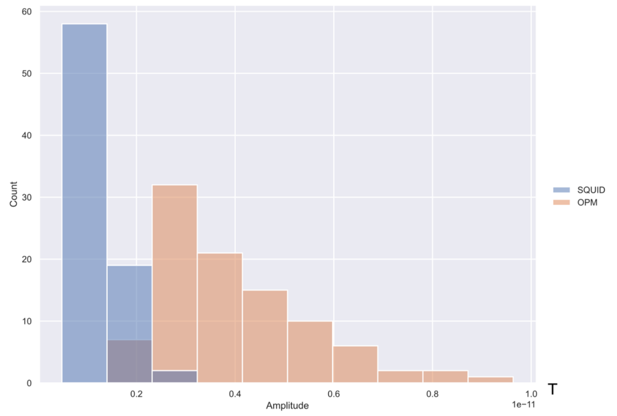

### Patient 11

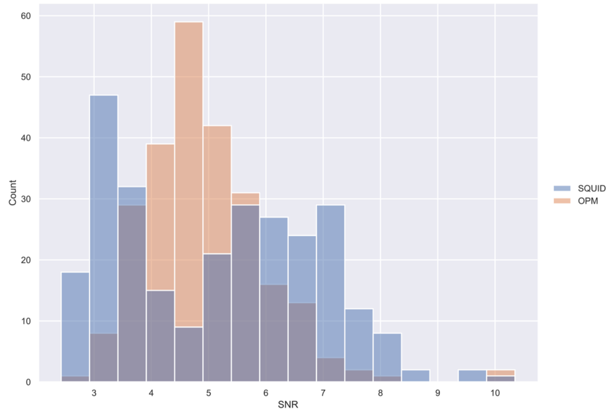

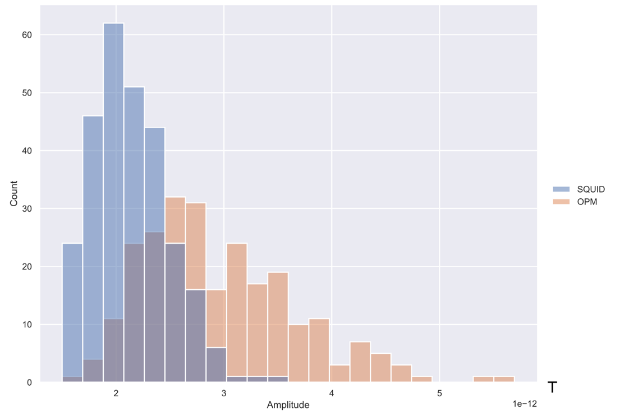

### Patient 20

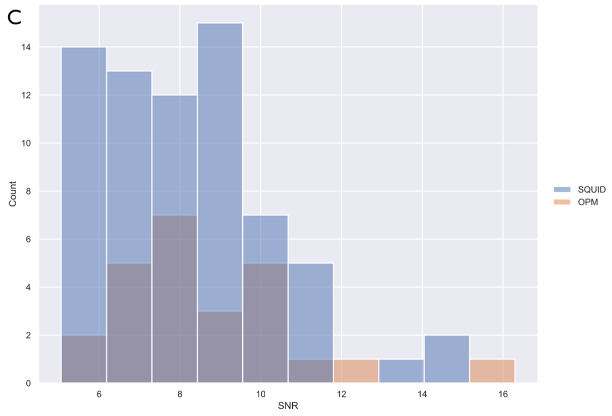

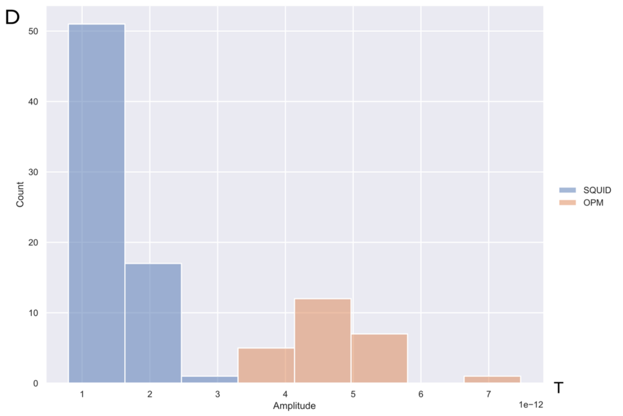

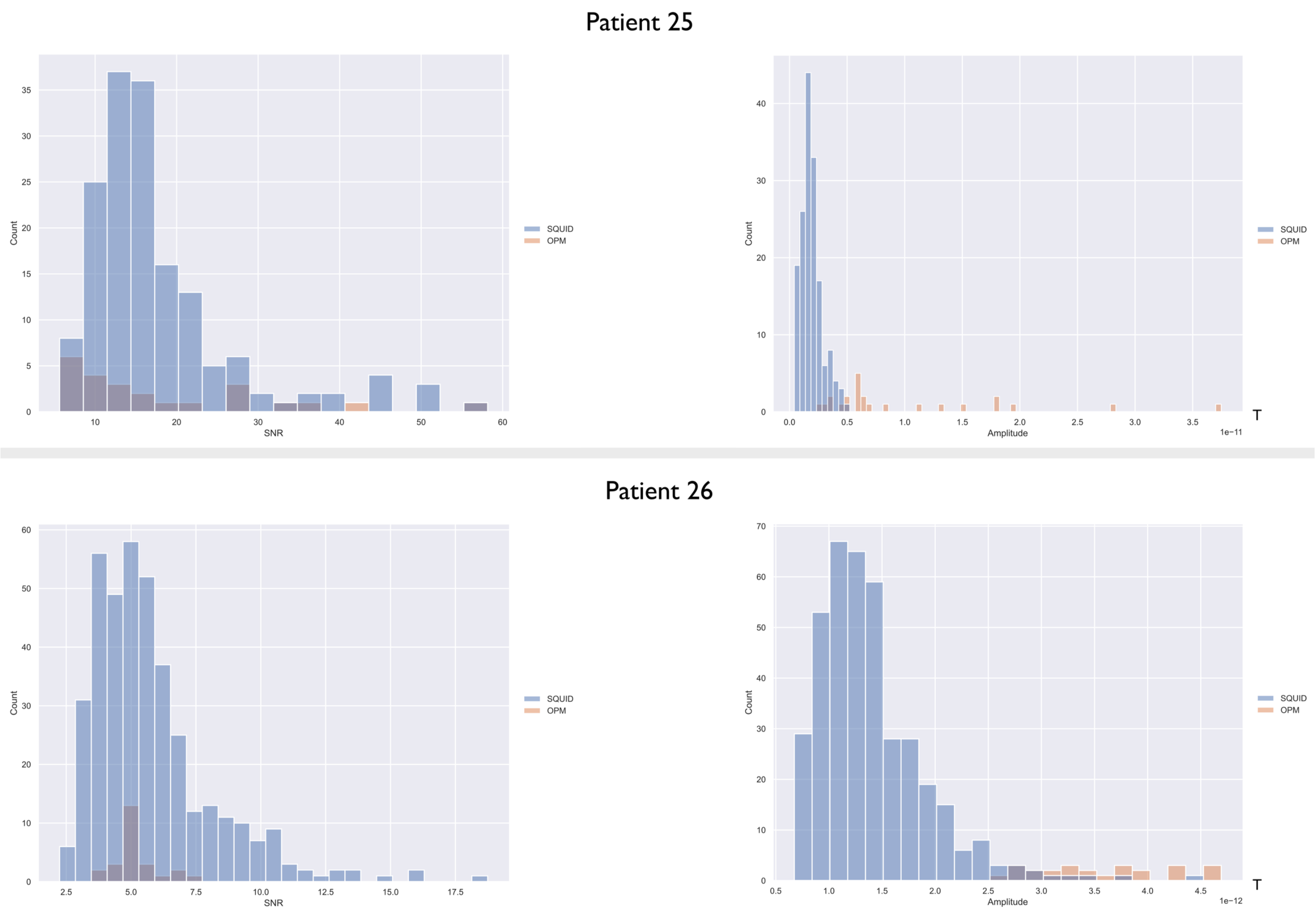

**Figure S2**: SNRs and maximum IEDs amplitudes for all patients – Left) Distribution of SNRs (SQUID-MEG in blue, and $^{4}He$ OPM-MEG in orange) – Right) Distribution of epileptic spikes maximum amplitude (SQUID-MEG in blue, and $^{4}He$ OPM-MEG in orange).

### Patient 3

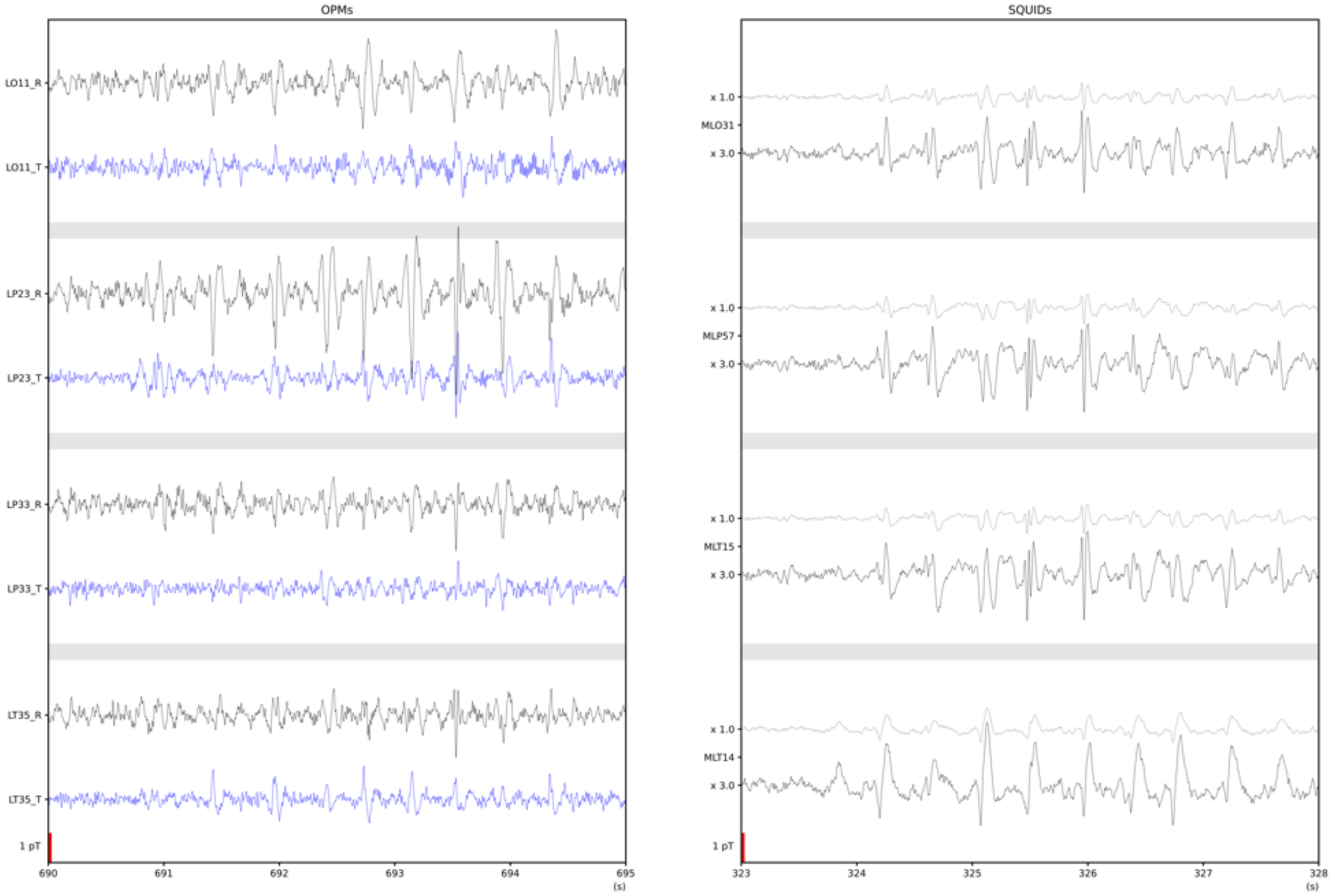

### Patient 11

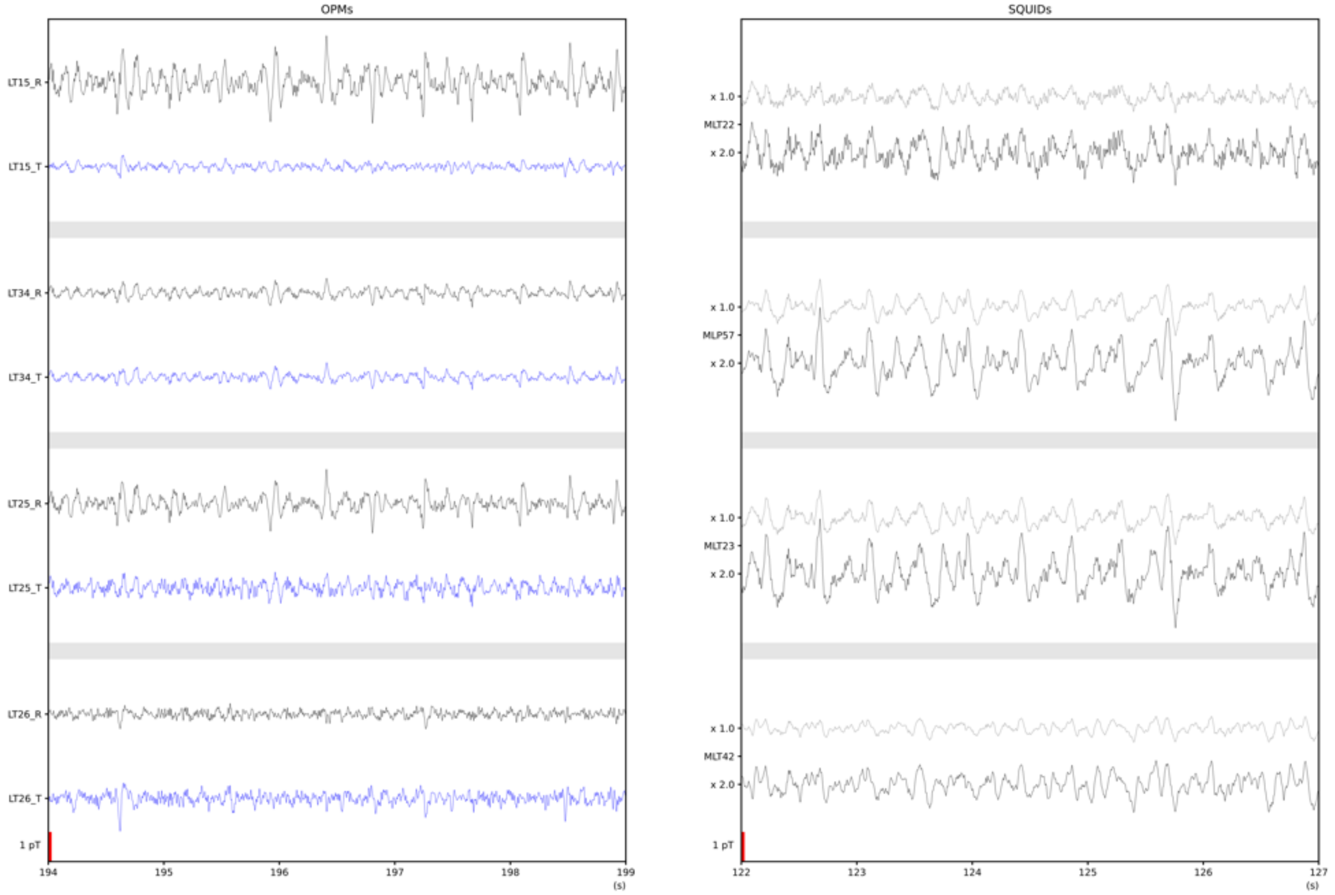

### Patient 20

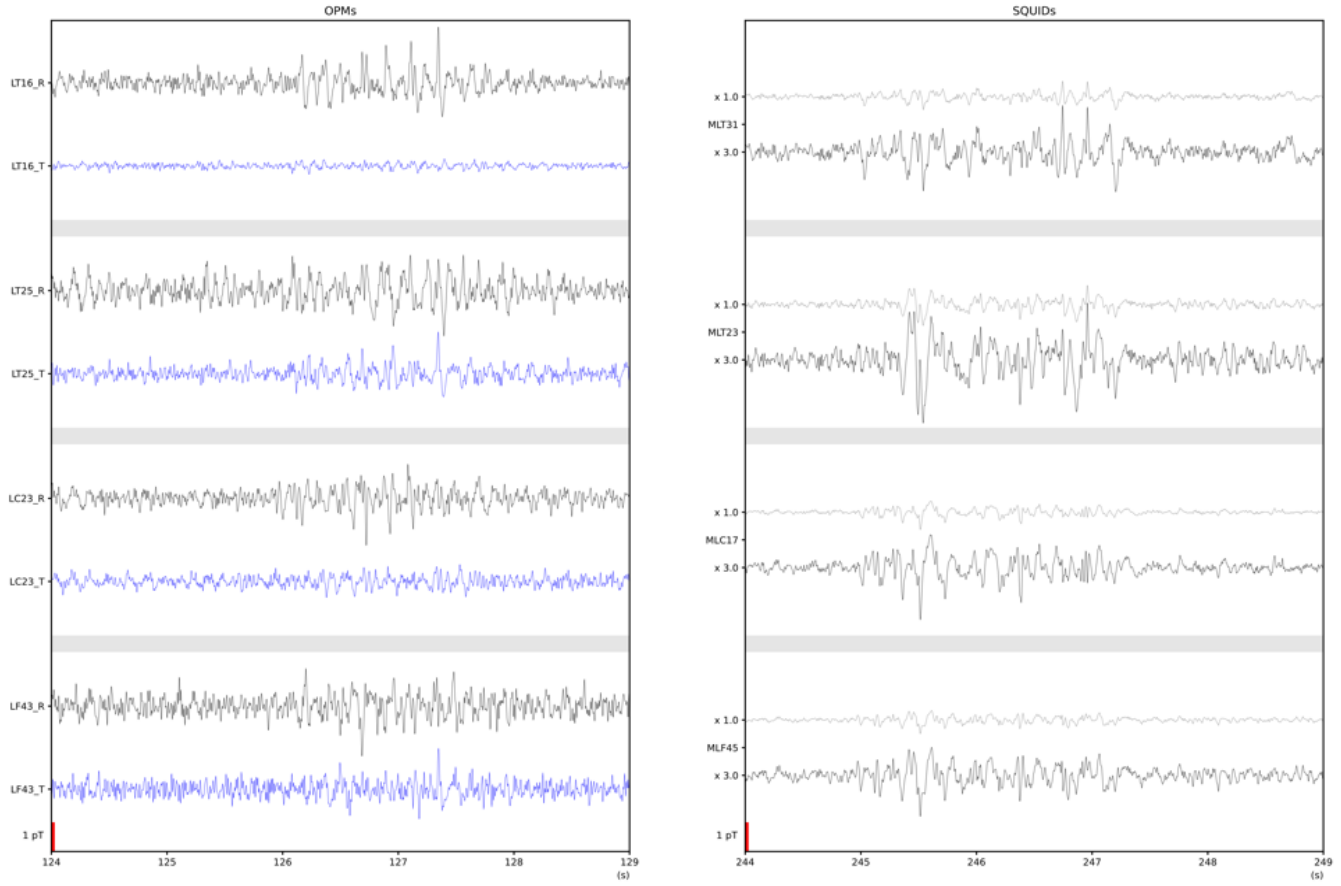

### Patient 25

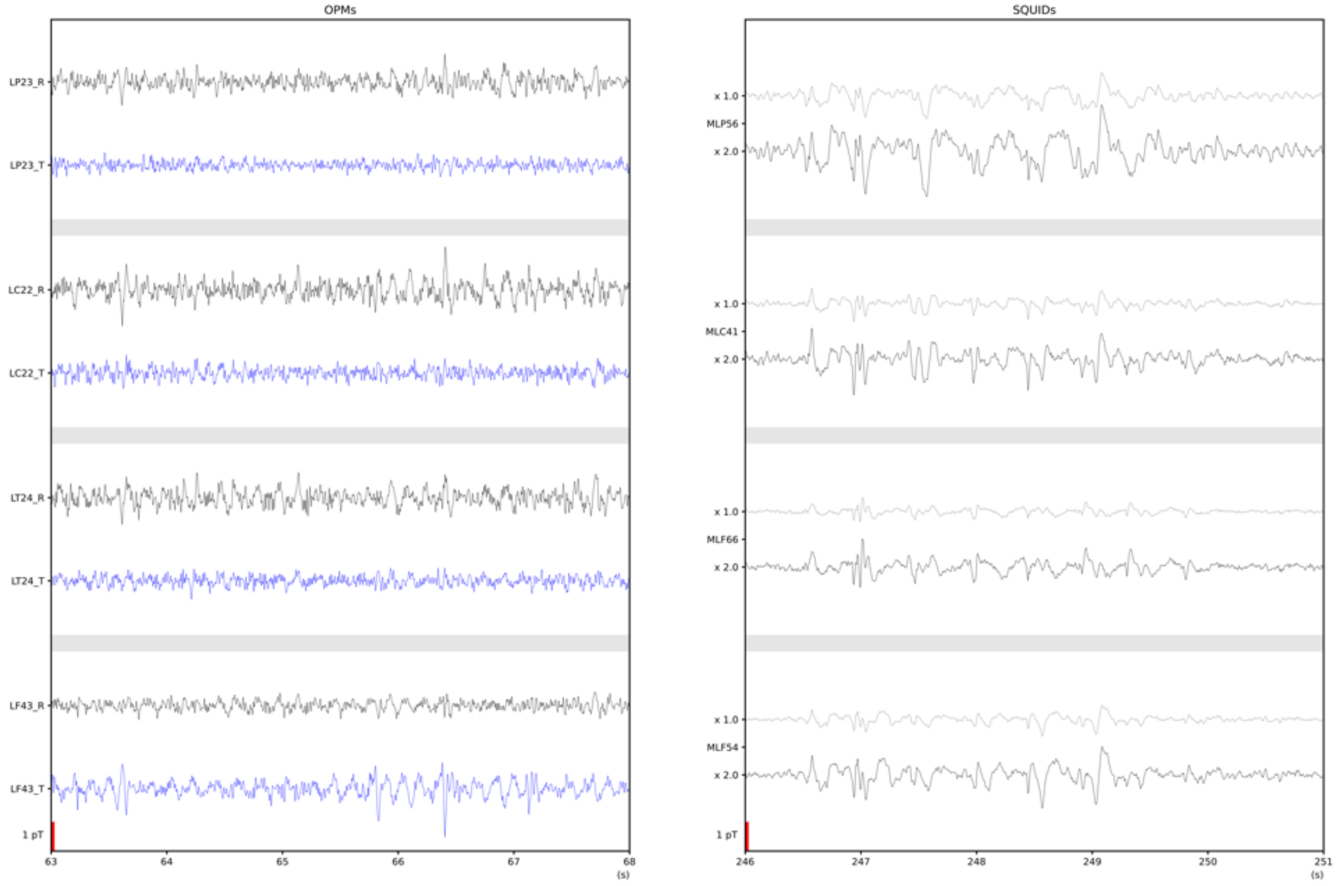

#### Patient 26

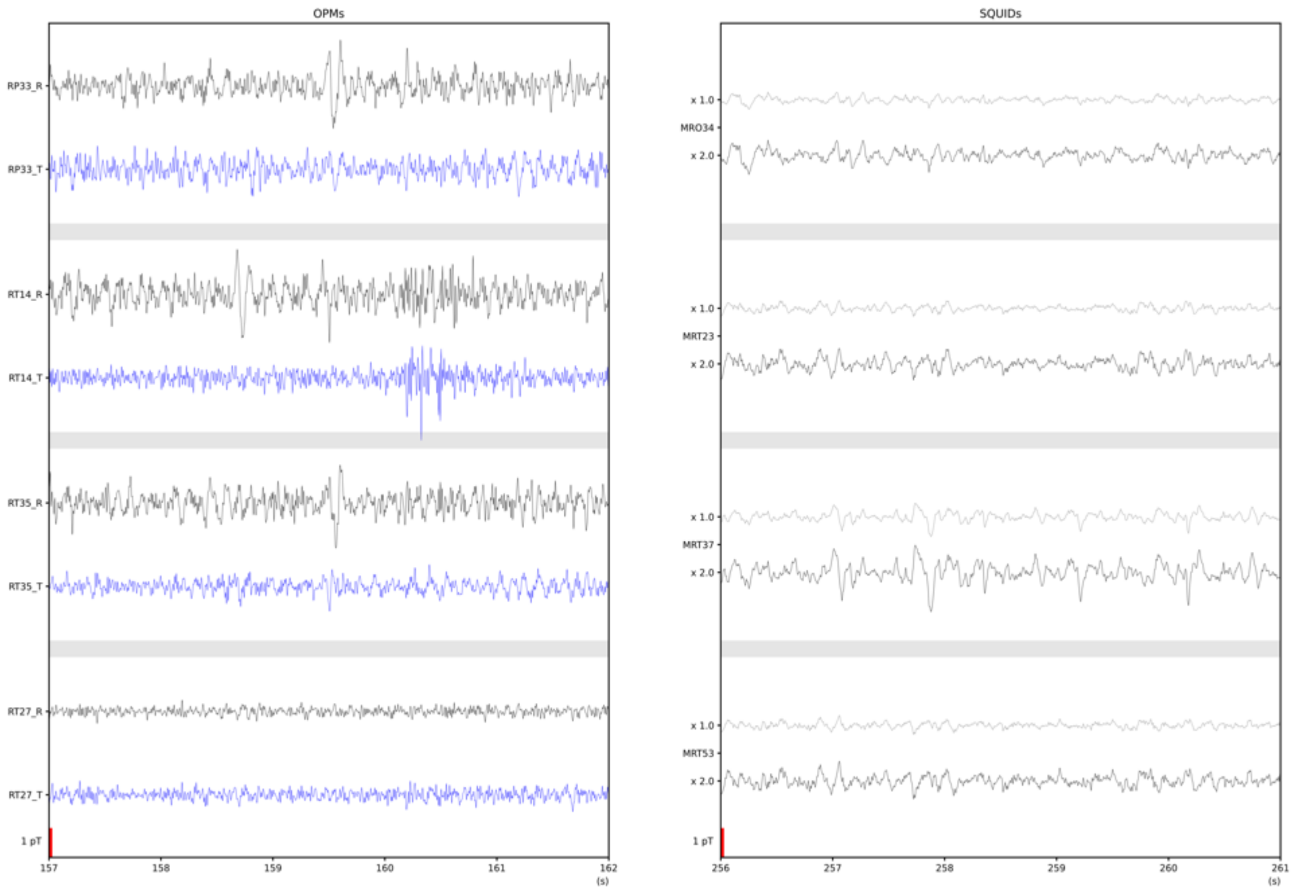

**Figure S3**: IEDs for all patients– For each patient the left panel shows the $^4$He OPMs traces with for each sensor in black the radial axis and in blue the tangential axis. The right panel shows the closest SQUID channel for each $^4$He OPM sensors with, in light grey, the trace at the same scaling as the $^4$He OPMs traces and in black the SQUID traces scaled for better comparison.

### Supporting Information: Tables

| | Sensor 1 | mean 1 | std 1 | Sensor 2 | mean 2 | std 2 | T value | Effect size | p value after bf correction |
|---|---|---|---|---|---|---|---|---|---|
| **Group level** | SQUID | 8,37 | 6,54 | OPM | 6,72 | 4,86 | 5,19 | 0,27 | 0,0040 |
| **Group level** | OPM tangential | 4,12 | 2,36 | OPM radial | 4,42 | 2,68 | -3,48 | -0,12 | 0,0040 |
| **Patient 26** | SQUID | 5,77 | 2,36 | OPM | 5,17 | 0,88 | 2,74 | 0,26 | 0,0999 |
| **Patient 25** | SQUID | 17,87 | 9,45 | OPM | 18,27 | 13,26 | -0,14 | -0,04 | 3,6044 |
| **Patient 11** | SQUID | 5,15 | 1,72 | OPM | 4,93 | 1,06 | 1,81 | 0,15 | 0,2557 |
| **Patient 20** | SQUID | 8,18 | 2,21 | OPM | 8,60 | 2,20 | -0,80 | -0,19 | 1,7742 |
| **Patient 3** | SQUID | 13,26 | 5,08 | OPM | 8,37 | 2,87 | 7,57 | 1,21 | 0,0040 |

**Table S1**: SNR detailed statistical results – Group level and individual level

| | Sensor 1 | mean 1 (T) | std 1 (T) | sensor 2 | mean 2 (T) | std 2 (T) | T value | Effect size | p value after bf correction |
|---|---|---|---|---|---|---|---|---|---|
| **Group level** | SQUID | 1,668E-12 | 6,6333E-13 | OPM | 3,7694E-12 | 2,8117E-12 | -15,06 | -1,28 | 0,0040 |
| **Group level** | OPM tangential | 1,4826E-12 | 1,1726E-12 | OPM tangential | 2,2858E-12 | 1,8543E-12 | -14,95 | -0,52 | 0,0040 |
| **Pat. 26** | SQUID | 1,3988E-12 | 5,0975E-13 | OPM | 3,5787E-12 | 6,1941E-13 | -16,89 | -4,21 | 0,0040 |
| **Pat. 25** | SQUID | 1,9230E-12 | 9,2967E-13 | OPM | 1,0226E-11 | 8,4865E-12 | -4,69 | -2,60 | 0,0040 |
| **Pat. 11** | SQUID | 2,1370E-12 | 3,4691E-13 | OPM | 2,9603E-12 | 7,0841E-13 | -16,54 | -1,50 | 0,0040 |
| **Pat. 20** | SQUID | 1,3309E-12 | 3,8609E-13 | OPM | 4,7172E-12 | 7,3829E-13 | -21,46 | -6,64 | 0,0040 |
| **Pat. 3** | SQUID | 1,1425E-12 | 4,4884E-13 | OPM | 4,0396E-12 | 1,5423E-12 | -17,43 | -2,44 | 0,0040 |

**Table S2**: IEDs maximum amplitude detail statistical results - Group level and individual level
